# RoMIA: A Framework for Creating Robust Medical Imaging AI

**DOI:** 10.1101/2023.04.10.23288377

**Authors:** Aditi Anand, Sarada Krithivasan, Kaushik Roy

## Abstract

Artificial Intelligence (AI) methods, particularly Artificial Neural Networks (ANNs) have shown great promise in a range of medical imaging tasks. Despite their promise, the susceptibility of ANNs to produce erroneous outputs under the presence of input noise, variations, or adversarial attacks is of great concern and one of the largest challenges to adoption in medical settings. Towards addressing this challenge, we explore the robustness of ANNs trained for chest radiograph classification under a range of perturbations reflective of clinical settings. We propose RoMIA, a framework for the creation of <u>Ro</u>bust <u>M</u>edical Imaging <u>A</u>NNs. RoMIA adds three key steps to the model training and deployment flow: (i) Noise-added training, wherein a part of the training data is synthetically transformed to represent common noise sources, (ii) Fine-tuning with input mixing, in which the model is refined with inputs formed by mixing data from the original training set with a small number of images from a different source, and (iii) DCT-based denoising, which removes a fraction of high-frequency components of each image before applying the model to classify it. We applied RoMIA to create six different robust ANNs for classifying chest radiographs using the CheXpert dataset. We evaluated the models on the CheXphoto dataset, consisting of naturally and synthetically perturbed images intended to evaluate robustness. Models produced by RoMIA show 3-5% improvement in robust accuracy, suggesting that the proposed methods can be a useful step towards enabling the adoption of ANNs in medical imaging applications.

## 1 Introduction

Artificial Intelligence is transforming the field of medicine in many ways, with applications spanning from drug discovery to genomics and, most prominently, radiology. Since AI has been particularly successful in computer vision, one of its most promising applications is to medical imaging. Artificial neural networks (ANNs), which are the class of AI models most frequently applied to computer vision tasks, have already been trained to diagnose various conditions from medical images, such as diabetic retinopathy [1][2], breast cancer and malignant lymph nodes from histopathological images [3], and pulmonary and cardiological conditions from chest radiographs [4]. The recent wave of promising research has led to significant interest in deploying these technologies in clinical settings. However, there are many hurdles that must be crossed before we can realize this potential.

Medical AI models, particularly ANNs, are first trained on a training dataset, and then tested in field trials before being deployed. One major challenge in this process arises from the differences between the data on which the models are trained and the data that they encounter after deployment[5]. Different usage scenarios of medical imaging ANN models can result in the introduction of noise and variations relative to the data that the model was trained on[6]. Since ANNs are known to be very brittle to input noise and variations[7], even ones that are imperceptible to humans[8], this poses a major roadblock to the deployment of successful medical imaging ANNs.

There are several scenarios where deployed medical ANNs encounter noise or variations that can impact the accuracy of their predictions. Since one promising area of deployment for medical imaging ANNs is telemedicine in remote areas that have a lack of trained physicians, images of scans are often sent through smartphone photos or messaging apps that distort and compress them [9]. Additionally, using imaging equipment made by different manufacturers or using different settings on the imaging equipment can create variations in the resulting images [10][11]. AI models have also demonstrated significant performance variation across different patient populations [12]. Inconsistency across any of these factors between the data a model in trained on and the data it is deployed on can result in an ANN making inaccurate predictions [13].

Recent work has demonstrated that variations and noise in the input can significantly reduce the accuracy of medical imaging ANN models [10][13]. Although there has been a large body of work in the AI community on improving the robustness of these models under noise and adversarial perturbations, very few efforts have focused on the medical domain. There are various unique challenges posed by the domain of medical imaging that make it essential to address robustness specifically in this context [13]. As described above, the nature of input noise and variations is primarily due to equipment differences, telemedicine, patient population; sources of variation seen in other settings (background objects, lighting, occlusion, etc.) are less relevant in medical settings [9] [10][12]. Furthermore, due to regulations and higher safeguards applied to medical data, adversarial attacks may be much less of a concern in this setting relative to other settings.

In this paper, we propose RoMIA, a framework to create more robust medical imaging ANN models that consists of three main steps: Noise-added Training, Fine-tuning with Input Mixing, and DCT-based denoising. In Noise-added Training, a fraction of the images in the training dataset are transformed by adding synthetic perturbations in order to make the trained model is more robust [14]. In Fine-tuning with Input Mixing, we fine-tune the trained model using a small amount of data from a different source in order to improve the model’s generalization [15]. Since only limited data from additional sources are likely to be available in practice, we use input mixing to avoid overfitting during this stage. Finally, in DCT-based denoising, we remove higher-frequency components in the input images before they are passed to the model for classification [16]. This is motivated by our observation that perturbations encountered in medical imaging settings largely impact the high-frequency components of the images.

We apply the RoMIA framework to the CheXpert dataset which contains 224,316 chest radiographs of 65,240 patients from Stanford Hospital [4]. We created a model that diagnoses Atelectasis, Cardiomegaly, Consolidation, Edema, and Pleural Effusion. We evaluated the model using the CheXphoto dataset, which consists of 10,507 natural photos and synthetic transformations of chest radiographs from 3000 patients [9]. For Fine-tuning with Input Mixing, we additionally used only 500 images from the ChestX-ray8 dataset from NIH [17]. Our experiments indicate that a baseline model trained on the CheXpert dataset has an Area Under Receiving Operating Characteristic (AUROC) drop of 10 to 14 % when evaluated on the CheXphoto dataset. The proposed framework improves AUROC by up to 5 %, underscoring its potential to create more robust medical imaging models.

## 2 Materials and Methods

In this section, we first describe the general process for training medical imaging ANNs, and the pitfalls of this process when it comes to creating robust models. We then present the RoMIA framework to increase model robustness and the methodology used to evaluate it.

### 2.1 Pitfalls in Current Training Methods

Typically, the creation of a medical imaging model starts with the collection of a large training dataset with training labels provided by physicians. In some cases, this may require years of data collection. For example, the CheXpert dataset of chest radiographs represents data collected over a period of 15 years [4]. Next, a deep neural network is either trained from scratch or a model trained on a different computer vision dataset such as ImageNet [18] is transferred and fine-tuned using the collected data. The model may be evaluated on held-out or entirely different datasets, and then deployed. Due to the difficulty of data collection, it is very challenging to evaluate the models on a wide range of datasets representing the entire swath of input conditions encountered in the field. Thus, when deployed, the model may be applied to data that contains noise or was captured by different equipment with different settings than encountered during model creation. Frequently, this leads to significant degradation in model performance [13].

### 2.2 Proposed Framework (change this heading)

Figure 1 describes the RoMIA framework to train more robust medical imaging models. We modify the standard model creation flow by adding three main components: Noise-added Training, Fine-tuning with Input Mixing, and DCT-based denoising.

**Figure 1:**
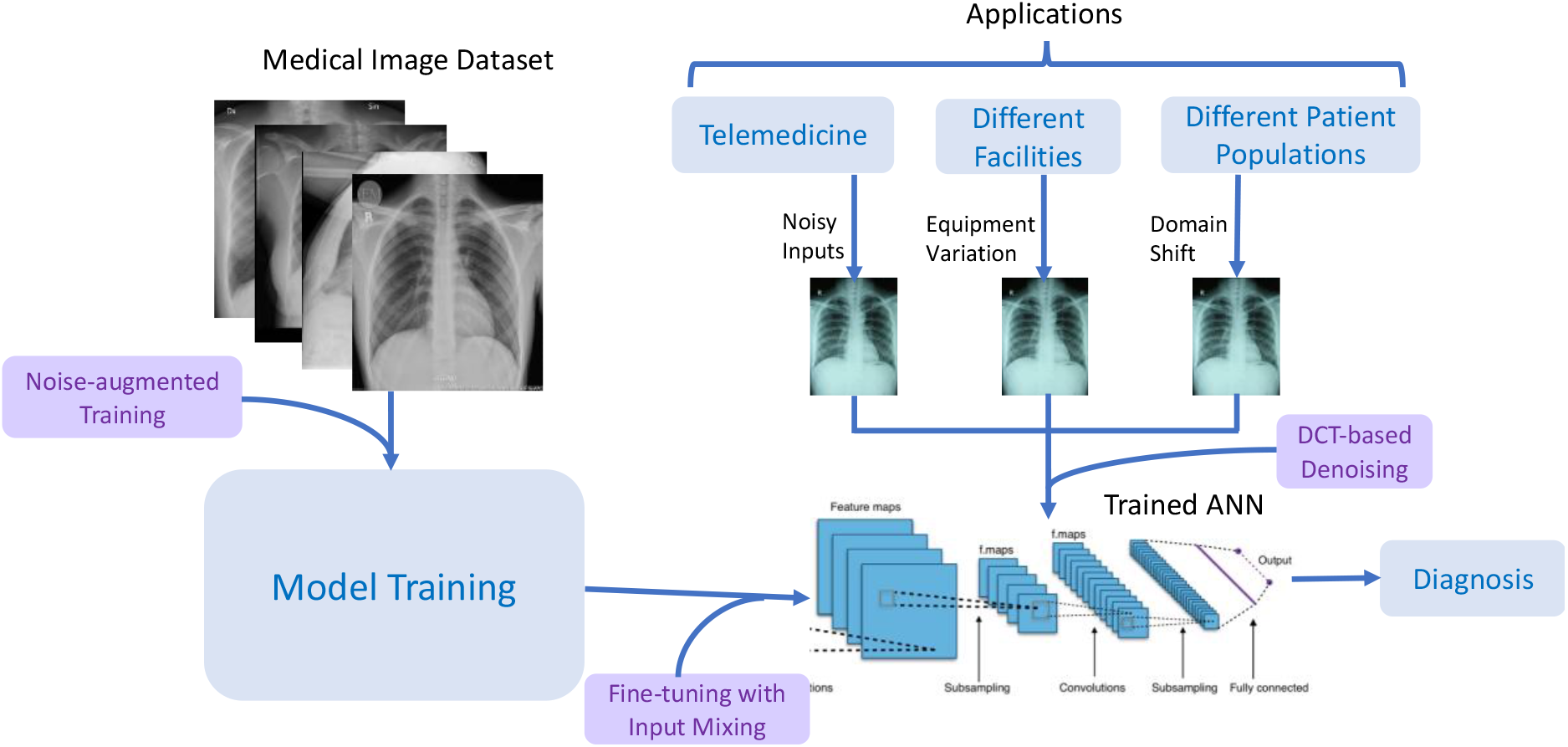
An Overview of the RoMIA Framework

#### 2.2.1 Noise-added Training

In Noise-added Training, we introduce synthetic perturbations into the training data that mimic those observed in medical settings. Following the approach of CheXphoto [9], we applied the following transformations:

- *Glare matte:* A filter designed to emulate the effect of glare observed when displaying the image on a matte screen.
- *Moire:* A filter designed to simulate the Moire effect, which produces repetitive interference pattern such as lines or stripes on the image due to limited resolution.
- *Tilt:* This transformation simulates a change in perspective that could result when a photograph of a medical image is taken using a device such as a smartphone [11].

Although we evaluated other transformations, such as changing brightness or contrast, or adding blur, we found that the three transformations considered above were the most effective in creating more robust models. We also consider two types of dataset transformations in which a specific percentage of the images in the dataset are transformed and either added (thereby expanding the dataset) or replace their original versions (thereby preserving the size of the dataset). We refer to these strategies as augmentation and replacement, respectively. All other training hyperparameters (learning rate, batch size, optimizer, epochs, etc.) were kept unchanged.

#### 2.2.2 Fine-tuning with Input Mixing

In Fine-tuning with Input Mixing, we fine tune the model with a very small amount of data from a different source to improve the model’s generalization ability. Since acquiring large amounts of additional training data may be challenging in practice, we limited ourselves to just 500 images, which correspond to around 0.22% of the original training set. For our experiments, we draw these images at random from the ChestX-ray8 dataset from NIH [17]. One challenge with using a very limited amount of data is that it could easily lead to overfitting. In order to prevent this, we use input mixing, a well-known technique where two images are combined into a composite input that contains information from both. Minimizing loss on mixed inputs has been shown to approximately correspond to maximizing robust accuracy [20]. We mixed the additional data with images from the original training set for the fine-tuning phase. We considered two different mixing strategies that have been proposed in the literature. With CutMix [15], a randomly selected patch of one input image is placed into another. With MixUp, the pixels of two images are averaged in a weighted manner to construct a composite image. In both cases, the labels from the two images being mixed are also combined to derive the target label for the composite input [20]. We mix the 500 images from ChextX-ray8 with 1000 randomly selected images from the CheXpert training set and fine-tune the model for 3 epochs with these mixed inputs. All other hyperparameters such as the learning rate and optimizer were the same as those used in the training stage.

#### 2.2.3 DCT-based Denoising

DCT-based denoising is based on the insight that most sources of noise disproportionately affect the high-frequency components of an image [21]. This is shown in Figure 2, which plots the percent difference in the top and bottom 1% of frequencies of the original and noisy images from the CheXpert [4] and CheXphoto [9] datasets, where the noisy images were produced using synthetic digital perturbations, synthetic photographic perturbations, and photos taken of the images with a smartphone camera. During inference, we add a preprocessing stage to the model which uses DCT (discrete cosine transform) to transform the image into the frequency domain, then removes a set percentage of high-frequency components, and finally computes the inverse DCT [16]. The percentage of high-frequency components to be removed from an image (denoted by η) is determined through an experiment where a small fraction of the training set (CheXpert, in our experiments) is subject to DCT-based denoising for different values of η. For each ANN, the largest value of η (which corresponds to the most aggressive denoising) that keeps the AUROC to within 0.005 of the original accuracy (where η = 0) is chosen.

**Figure 2:**
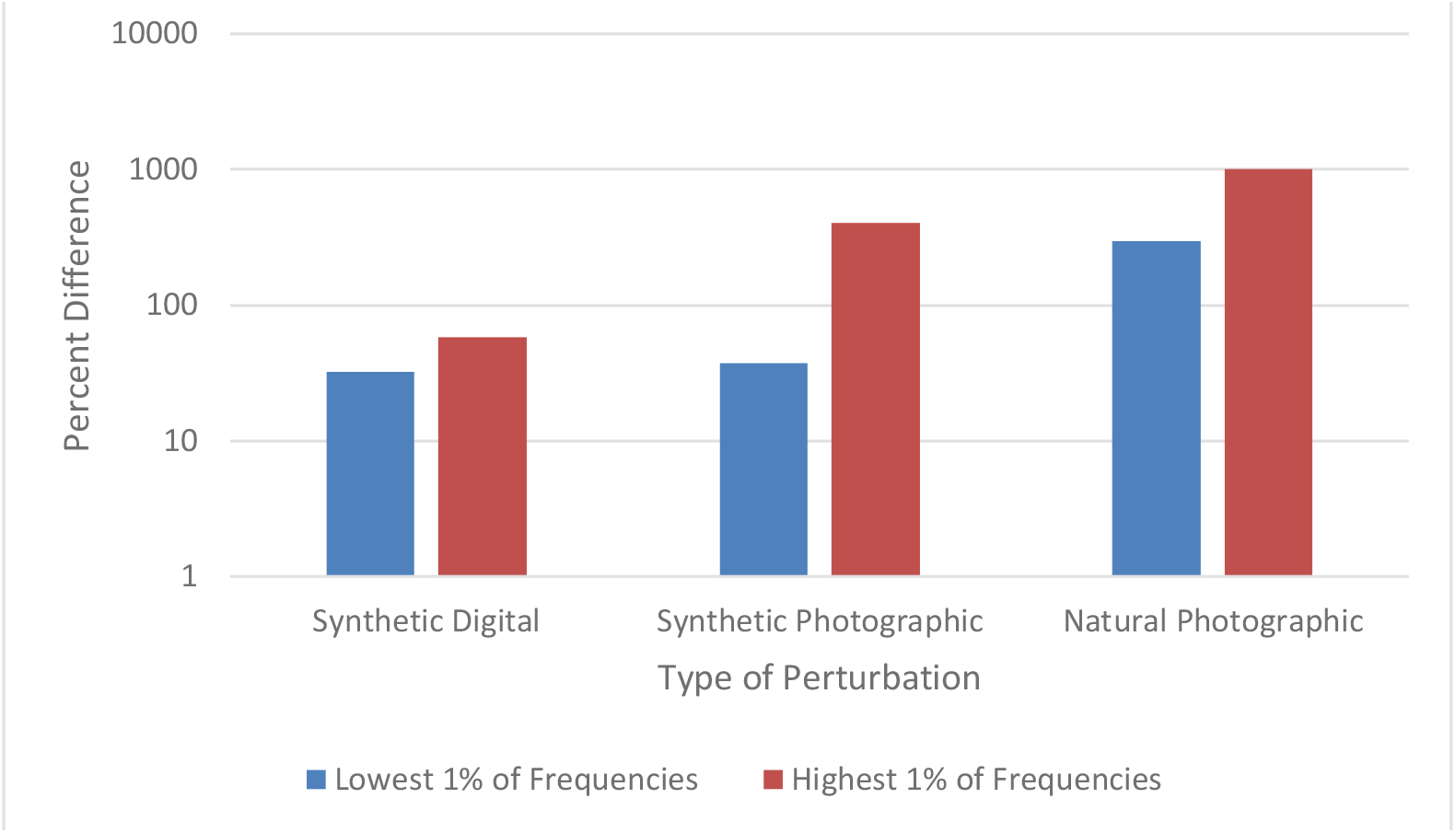
Percent Difference Between CheXpert and CheXphoto in Top and Bottom 1% of Frequencies

To summarize, the proposed flow to create robust medical imaging ANNs consists of transferring a model trained on ImageNet to the target medical imaging dataset using noise-added learning, then fine-tuning the resulting model with input mixing, then finally adding a DCT-based denoiser to the model before deployment.

### 2.3 Experimental Setup

We implemented the RoMIA framework using the PyTorch [22], TensorFlow [23], libAUC [24], and OpenCV [25] libraries. We applied the framework to create models for classification of chest radiographs. The base models were selected from popular image classification networks trained on the ImageNet [18] dataset (see Table 1). We specifically created a model to detect Atelectasis, Cardiomegaly, Consolidation, Edema, and Pleural Effusion. Accordingly, the final fully connected layer of each base model was removed and replaced with a layer with five outputs. These models were then transferred using the CheXpert [4] dataset, which contains 224,316 chest radiographs of 65,240 patients from Stanford Hospital. For the fine-tuning step, we randomly selected 500 images from NIH’s ChestX-ray8 [17] dataset. For the MixUp [20] strategy, we use a beta distribution to select values between 0.4 and 0.6 to determine λ, the image mixing ratio. We evaluated the models on the CheXphoto [9] dataset, which consists of 10,507 natural photos and synthetic transformations of chest radiographs from 3000 patients.

**Table 1:** Baseline Model Characteristics

| Model | Parameters | ImageNet Accuracy |  | CheXphoto<br>Baseline AUROC |
| --- | --- | --- | --- | --- |
|  |  | Top-1 | Top-5 |  |
| DenseNet121 | 8.0M | 74.434 | 91.972 | 0.7651 |
| DenseNet169 | 14.1M | 75.6 | 92.806 | 0.7691 |
| DenseNet201 | 20.0M | 76.896 | 93.37 | 0.7677 |
| ResNet18 | 11.7M | 69.57 | 89.24 | 0.7789 |
| ResNet34 | 21.8M | 73.27 | 91.26 | 0.8025 |
| ResNet50 | 25.6M | 75.99 | 92.98 | 0.7905 |

## 3 Results

In this section, we present results from evaluation of models created using the RoMIA framework. We first present the difference in AUROC of the baseline models when evaluated on a subset CheXpert and CheXphoto images. Next, we present the performance of models trained using RoMIA and compare them to the baseline models. Subsequently, we perform an ablation study to investigate the contribution of each of the three components (Noise-added learning, Fine-tuning with input mixing, DCT-based denoising) to the overall improvement in robustness. We then explore different dataset transformation techniques for the Noise-added Training step and evaluate their impact on the model performance. We also compare the performance between the CutMix and MixUp strategies for input mixing in the fine-tuning step. Finally, we explore the determination of the parameter η which controls the percent of high-frequency components removed from the input during DCT-based Denoising

### 3.1 Lack of Robustness in Baseline Models

A key motivation for this work is that baseline models trained on a certain dataset perform significantly worse on similar datasets with added noise. To demonstrate this in the context of CheXpert and CheXphoto, we study the differences in AUROC of a baseline model trained on CheXpert and then applied to both CheXpert and CheXphoto data. Figure 3 presents the AUROC scores for the baseline models on the CheXpert and CheXphoto data. The figure shows a degradation of 10-14% across ANN models, underscoring the need to create more robust models in the context of medical imaging.

**Figure 3:**
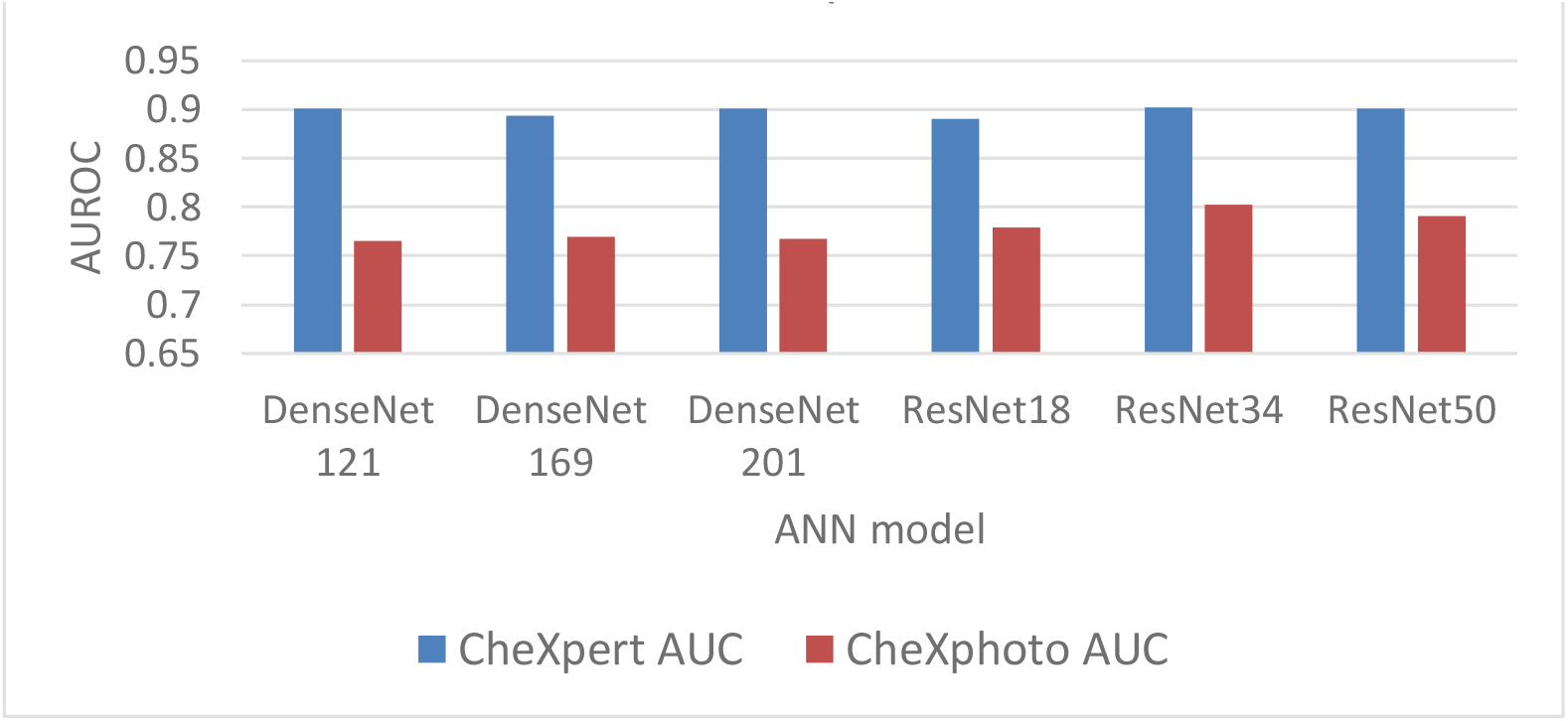
AUROC of Baseline Models on CheXpert and CheXphoto

### 3.2 Overall Improvements from RoMIA and Ablation Study

Our proposed framework consists of three techniques to improve robustness, so we conduct an ablation study to evaluate each component. Figure 4 shows the baseline accuracy, the results of the ablation study (applying each of the three techniques in RoMIA individually), and the resulting AUROC score when all three techniques are combined in RoMIA. To capture the benefits of the proposed framework, we first look solely at the CheXphoto AUROC values for the baseline and RoMIA models. We observe around 3-5 % improvement in AUROC, suggesting that the proposed framework is capable of creating substantially more robust models. We also observe a larger improvement in robustness on deeper models, such as ResNet50 and DenseNet201. We hypothesize that this is because deeper models can better learn the more diverse training data which they are presented in the RoMIA framework.

**Figure 4:**
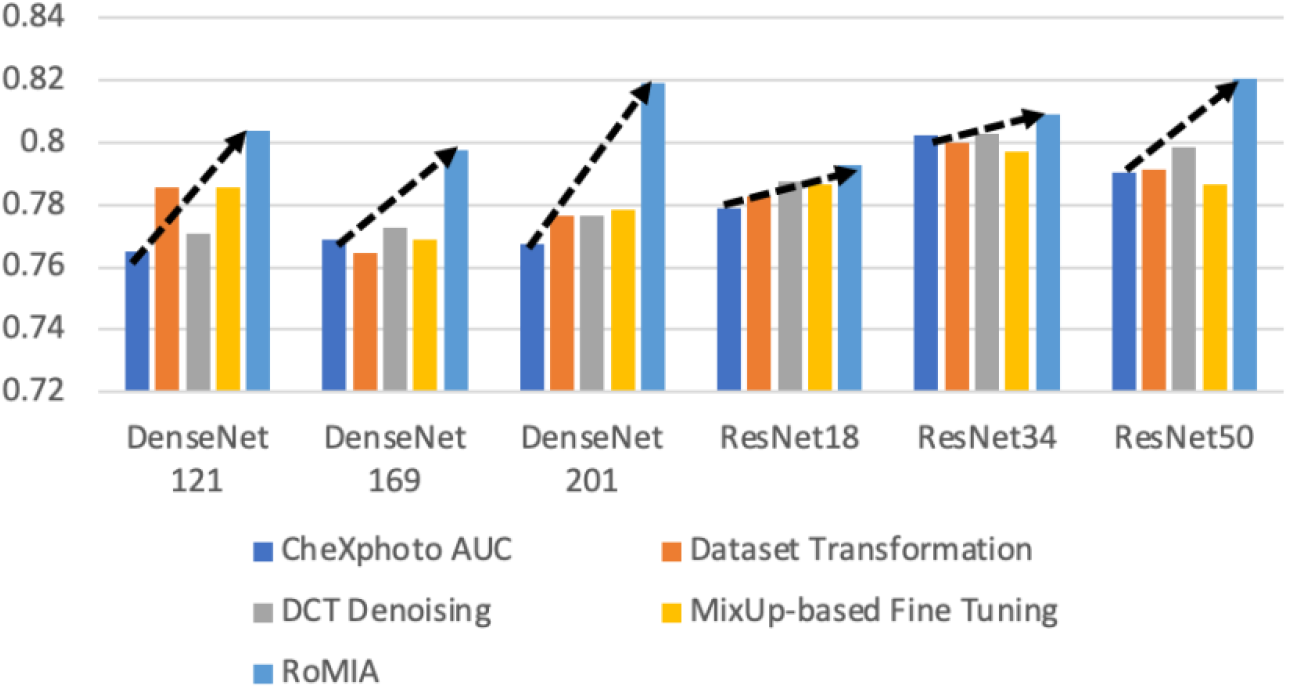
AUROC Improvement from Each Individual Technique and Combination (RoMIA)

Figure 4 also presents the results of our ablation study to evaluate each of the three components in the proposed framework. We do this by evaluating the CheXphoto AUROC when each technique is applied individually. We observe that overall, each technique has a positive impact on robustness. The combination of three techniques used in RoMIA boosts AUROC by up to 5%. We evaluate each technique in more detail in subsequent sub-sections.

### 3.3 Contributions from Noise-Added Training

Figure 5 explores the impact of various dataset transformation techniques used in noise-added learning. Specifically, we transformed 10%, 25%, and 50% of the training samples in the CheXpert dataset and either added them to the dataset (augmentation) or replaced the original samples with them (replacement). We observe that the 25% replacement strategy worked best across all networks. We note that this strategy does not impact training time, as the only overhead incurred is a one-time transformation (noise addition) to the inputs, which is insignificant.

**Figure 5:**
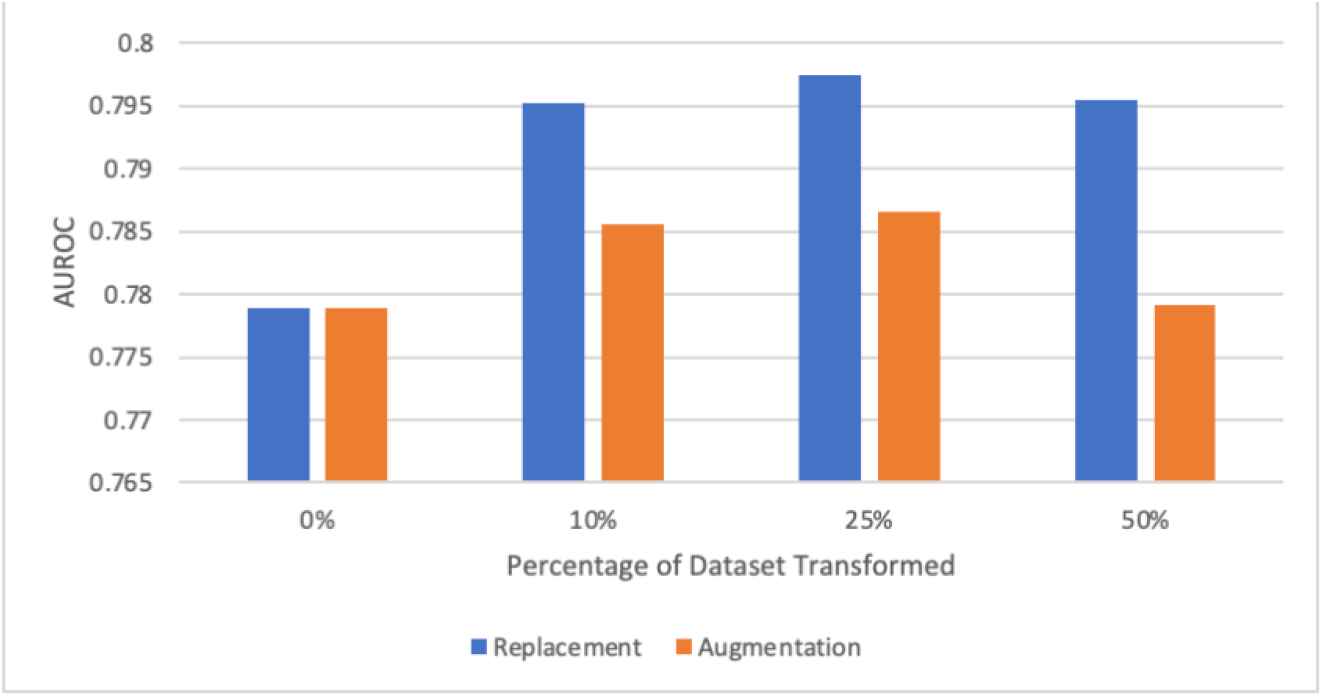
AUROC for Dataset Replacement and Augmentation in ResNet18

### 3.4 Effect of Fine-tuning with Input Mixing

Several approaches to input mixing have been developed. We focus on CutMix [15] and MixUp [20], the two most widely used strategies. To determine which strategy yields higher improvement in robustness, we compare the AUROC boosts on the CheXphoto dataset when each strategy is applied without other techniques to an ANN model. Figure 6 compares the two strategies in the Fine-tuning with Input Mixing step. We observe that the results from the two mixing strategies are similar, with MixUp providing slightly better results, which motivated our decision to use MixUp in the final RoMIA framework. CutMix and MixUp are both known to improve robustness, where minimizing loss on mixed inputs leads to better robust accuracy, since these strategies reduce overfitting and improve model generalization.

**Figure 6:**
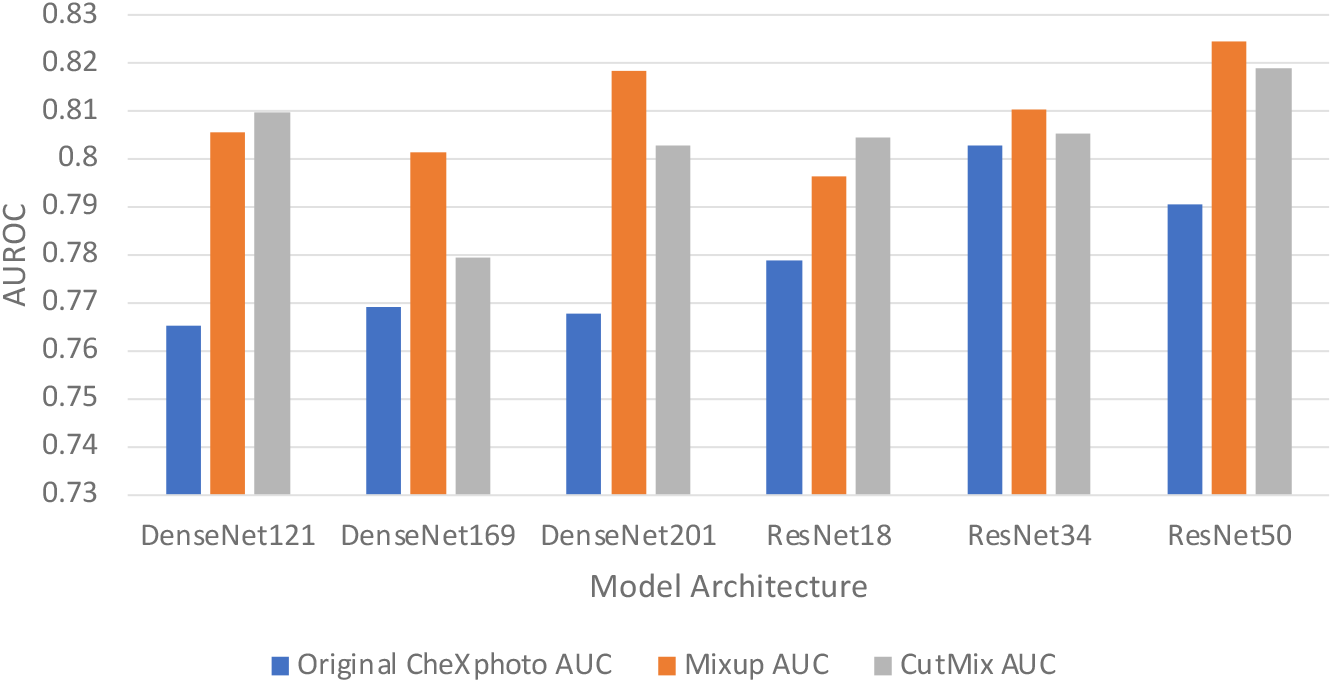
AUROC Improvements when using CutMix vs. MixUp based fine tuning

### 3.5 Selection of η in DCT-based Denoising

A key feature of our framework is the DCT-based Denoising step, which removes high-frequency noise from the inputs. We use the parameter η to denote the percentage of high-frequency components removed from each image. In Figure 7, we consider the impact of the choice of the parameter η by showing how different η values affect CheXpert and CheXphoto AUROC. Due to the nature of X-ray radiographs, we find that removing a large fraction of the high frequencies does not have a detrimental impact on performance for either dataset and on the other hand improves accuracy on the noisy (CheXphoto) data. We determine η as the largest value that results in a less than 0.5% decrease in accuracy on the clean (CheXpert) dataset. We observe that this value of η in fact improves performance on the CheXphoto dataset. This result also underscores the efficacy of DCT-based denoising.

**Figure 7A:**
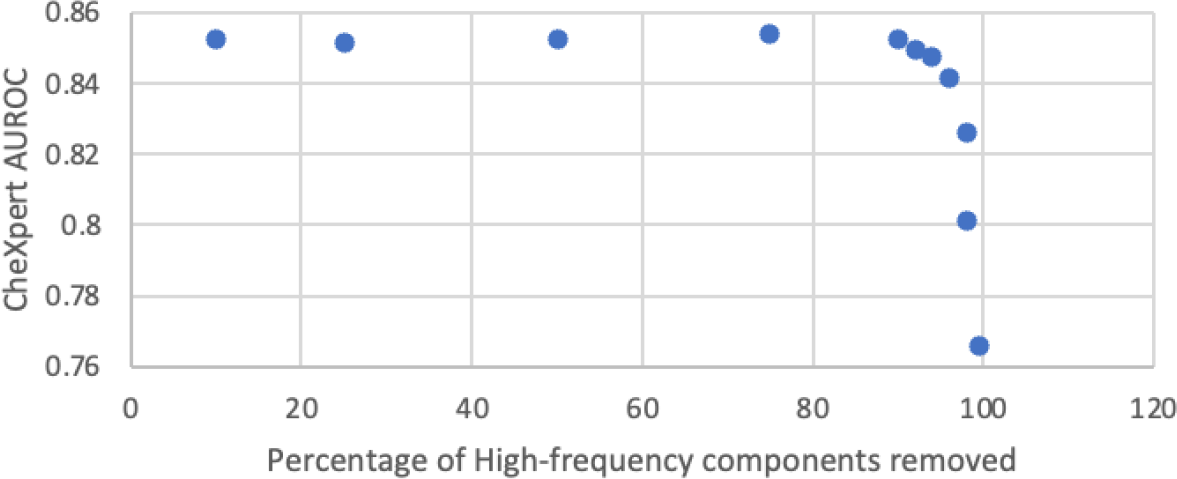
Determination of η based on CheXpert AUROC when high-frequency components are removed

**Figure 7B:**
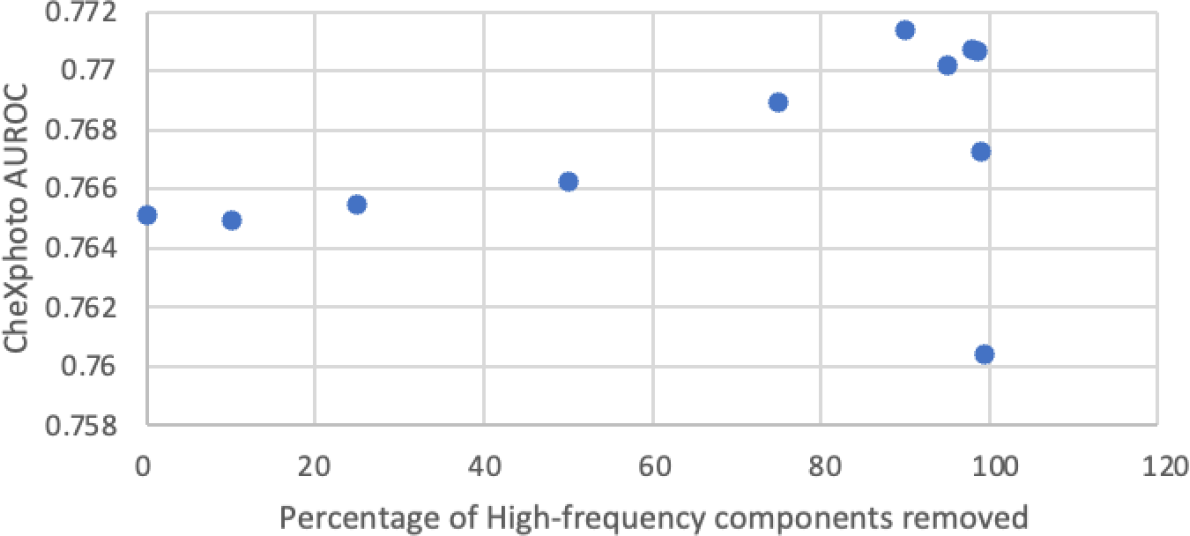
Effect of removing high-frequency components on CheXphoto AUROC

## 4 Discussion

The success of AI in recent years has led to significant interest in applying it to the medical field. In particular, since ANNs have been very successful in image processing applications, they are frequently being applied to medical imaging tasks. One of the challenges that must be addressed when applying AI to any critical application, and certainly to medical imaging, is their robustness under conditions encountered in the real world. Previous research has shown that ANNs can be very brittle in the presence of input noise and variations. Our work is a first step towards improving the robustness of medical imaging ANNs, with a particular focus on the kinds of noise encountered in medical settings. Although our proposed method focuses on ANNs classifying chest radiographs, the RoMIA framework can be applied to other domains in medical imaging.

### 4.1 Related Work

Several research efforts have explored the use of AI, and specifically, ANNs for medical imaging efforts. Building on these efforts, systems that support diagnosis are in various stages of deployment. These include systems for retinal scans [1][2][26], breast cancer detection [27], and skin cancer detection [28], among others. Given the limitations of ANNs when processing inputs with variations, noise, and adversarial perturbations, there have been several efforts that have pointed out these limitations. For example, it has been shown that chest radiographs with added variations reflecting those found in medical settings as well as photographs of medical images caused significant degradation in accuracy [9]. Additionally, ANN models trained on data from one hospital demonstrate considerably lower performance on data from a different hospital [10]. Adversarial perturbations have been shown to have a drastic impact on accuracy of ANNs used in oncology [29].

Given the above findings, it is critical to develop techniques to create more robust AI models. Very little work to date addresses this challenge. Adversarial training, where adversarial inputs are included in the training process, has been shown to improve the accuracy of ANNs used in medical imaging [29]. However, we are unaware of any work that improves robustness in the presence of noise sources more commonly observed in medical settings, such as equipment differences and the use of electronic scans and photographs in telemedicine. Our work is a step in this direction and shows that it is possible to create more robust AI models by addressing these noise sources during the model creation process.

### 4.2 Future Work

While the RoMIA framework achieves significant improvements in robust accuracy, there remains a gap in accuracy on clean and noisy inputs that could form a challenge for future work. One possible direction is to address robustness when training from scratch, in contrast to RoMIA, which only addresses it starting with the transfer learning step. Another interesting direction would be evaluating these techniques in a broader range of medical imaging applications. Given the criticality of medical imaging applications, robustness evaluation should be made a standard part of the regulatory evaluation process for these models. Finally, human checking of the output of AI models is one way of improving the confidence in their decisions. This could be enabled by creating explainable models that produce a human-interpretable justification for their decisions. Addressing these issues will go a long way towards enabling the adoption of AI-based medical imaging in clinical practice.

## Data Availability

All data produced in the present study are available upon reasonable request to the authors

